# A modification to the Maquet Flow-i anaesthesia machine for ICU ventilation

**DOI:** 10.1101/2020.04.06.20054882

**Authors:** Andrew J. Robinson, William London, Laszlo Kotan, Warwick Downing

## Abstract

The authors present an easily manufactured modification of Getinge Group’s Maquet Flow-i anaesthesia machine that gives it potential to be used long-term as an Intensive Care ventilator for emergency circumstances. There are some 7000 such machines in use worldwide, which could assist in increasing ICU ventilated bed capacity in a number of nations. The authors believe this modification has potential as a solution to increasing ventilator numbers for the COVID-19 pandemic, in hospitals where the Flow-i is underutilised for its designed purpose during this emergency. The technical drawing files are downloadable on the GrabCAD website and are Creative Commons (CC-BY 4.0) licensed to allow local manufacture of the modification. We welcome other Flow-i users and engineers to join us in troubleshooting this project on the associated GrabCAD discussion group during this ‘pre-print’ phase of research.

## Introduction

The COVID-19 pandemic has already overwhelmed Critical Care capacity in Italy and looks set to do the same in many other countries. If spread is not adequately contained, the critically ill cohort from this infection produces such a number of patients with Sudden Acute Respiratory Syndrome (SARS) in whom positive pressure ventilation is indicated, that ventilated bed capacity is rapidly overwhelmed, and patients are left untreated. Ventilated beds are in short supply worldwide, so access to additional ventilators of adequate quality to ventilate coronavirus SARS patients is urgently required. This pre-paper introduces a ‘proof of concept’ modification to the Maquet Flow-i anaesthesia machine (Getinge, 2020a) that makes it more suitable for use as an Intensive Care ventilator.

### COVID-19 and ventilated bed capacity

The SARS-CoV-2 infection COVID-19 has expanded worldwide since its lowly beginnings as a zoonotic infection in or near Wuhan, PRC (Andersen et al., 2020) sometime in late 2019. It is currently a WHO declared pandemic and wreaking havoc with the health and economic stability of the planet (WHO, 2020). Unchecked, it has the capability to cause millions of deaths worldwide. WHO and most local public health responses throughout the world hves concentrated on ‘flattening the epidemiological curve’. An exponential increase of SARSCoV-2 virus spread leads to unprecedented demand for critical care services. Flattening the curve is an attempt to reduce the mortality rate and avoid completely overwhelming healthcare systems. An anticipated side effect of ‘flattening the curve’ is, however, sustained demand for Intensive Care services extending over a long period of time for those nations unable to reduce spread early on. While the risk of an individual patient needing respiratory care can appear low on first examination, across a population COVID-19 produces a large number of such patients. These may require critical care for 14 days or more (Zhou et al., 2020). Demand for ventilators in the worst affected of these is thus predicted to be very high. Ventilated bed capacity worldwide is low. New Zealand has only some ∼200 ventilated beds nationwide under normal circumstances, and this number is barely adequate for normal demand. Other nations are in a similar state. However, Italy —the world epicenter of the disease as design commenced— had above average ICU beds/100 000 by European standards prior to the pandemic (Rhodes et al., 2012) and was still overwhelmed. A variety of stop-gap solutions to expand ventilator capacity are therefore required while manufacture of these devices is ramped up to meet demand. This paper proposes one such solution for hospitals which have Maquet Flow-i anaesthesia machines available.

### Maquet Flow-i

The Flow-i (Getinge, 2020a) is a modern anaesthesia machine designed for use in an Operating Room environment to provide short-term ventilation and delivery of volatile anaesthetic gases to patients undergoing general anaesthesia. There are some 7000 such units installed worldwide. The machine is based in part on ventilator technology that Getinge Group purchased from Siemens and is substantially similar in function to the Maquet Servo-i Intensive Care ventilator (Getinge, 2020b). Modern intensive care ventilators are ‘total loss’ systems, in that the gases supplied to the breathing system are filtered through a High Efficiency Particulate Air (HEPA) filter then passively expelled to atmosphere after use. The Flow-i anaesthesia machine, however (and in common with other such machines), is designed to allow the expiratory gases to be recirculated with CO_2_ scavenging in ‘low flow’ mode with minimal losses to atmosphere. This ‘partial loss’ system allows preservation of expensive and environmentally damaging volatile anaesthetic gases and lowers consumption of piped oxygen and air. It does this by directing all expired air internally through a *Volume Reflector*, a convoluted reservoir that sits under the anaesthetist’s writing surface that collects the patient’s expired air for recirculation. The Volume reflector is connected to the *Patient Cassette* by the *Socket*, a pair of linking tubes and silicone seals to prevent leaks. The Patient Cassette is the insert which contains the circle non-return valves and interfaces with the ventilator mechanisms, volatile delivery system, Adjustable Pressure Limiting valve (APL), and internal gas bench. All three of these inserts are removable and sterilizable.

### Investigating longterm ventilation options on an unmodified Flow-i

Anaesthetic machines are designed to be used for each patient for only hours rather than the days or weeks required of an ICU ventilator, and clinical experience suggests that they tend to experience more condensation and rainout than machines purpose built for ICU. Experience of using such machines for longer term in a pandemic situation is absent from the literature but will no doubt become more available as the pandemic progresses. While anaesthesia machines are able to run for short periods of time without scavenging, they are designed to be used with a constant low-level vacuum applied via the Anaesthesia Gas Scavenging (AGS) port and in a positive pressure Operating Room environment.

#### Low flow not feasible for ICU

When ventilating patients for longer procedures or in an emergency and in place of an ICU ventilator, it is generally advised to run on low flows to maximise humidification. However, the first author’s department has four years of experience of using the Flow-i under low flow conditions for long procedures. There is generally considerable rainout in the breathing system which requires decanting from the circuit every few hours if water buildup is not to interfere with ventilation. When wet Heat and Moisture Exchangers (HME) are less efficient at heating inspired gases, due to the energy lost to vaporizing excess water with each breath. This makes Flow-i low flow undesirable for longer term use in COVID-19, as decanting will require loss of Positive End Expiratory Pressure (PEEP) and increase potential for aerosol contamination with viral particles. Low flow also requires the use of soda-lime to scrub CO_2_ from the breathing gas, which adds another expense and a resource that might be depleted.

#### Total Loss undesirable long-term with HME alone

In order to avoid the circuit rainout issue, the machine could be used in high flow modeand used as a total loss system like an ICU ventilator by setting the Fresh Gas Flow (FGF) above Minute Volume and thus allowing all unused air, oxygen, and carbon dioxide from cellular respiration to be vented to atmosphere or AGS. This renders the HME more efficient in heating, but less so for humidification. Reduced humidification risks drying patient mucosa, ciliary damage, thickened secretions, atelectasis (Branson, 2009), risks of poorer outcomes (Misset et al., 1991), and a higher incidence of artificial airway blockage. While a Cochrane review of the available literature suggests there is not a major difference in ICU outcomes between HME and active humidification, it did so with the caveat “HMEs may not be suitable for patients with limited respiratory reserve or prone to airway blockage” (Kelly et al., 2010), something which certainly pertains in the COVID-19 ARDS cohort of patients. Our ICU uses Fisher & Paykel (F&P) active humidification for patients ventilated for longer than 24 hours, so the HME alone option was considered undesirable.

#### Total Loss with active humidification too wet

When our normal ICU ventilators are used with F&P active gas heating and humidification, we do not suffer significant rainout in the patient circuit. The Flow-i was therefore set up with an appropriate circuit and allowed to operate as a total loss system over night. This was done once without scavenging (AGS), and on the second night with AGS. In the AGS condition, the breathing system remained clear of liquid water, but condensation formed throughout the Volume Reflector. Without AGS, it was possible to decant liquid water from the Volume Reflector after a mere 12 hours.

It was concluded that use of a Flow-i with active humidification would move the rainout problem from the breathing system, but result in water pooling in the Volume Reflector during the prolonged ventilation required for a COVID-19 SARS patient. This would be particularly so if used in an ICU environment without AGS. Accumulated water would progressively interfere with ventilation dynamics and, at some point, require the Patient Cassette, Socket, and Volume Reflector be re-processed and dried out. The interruption to ventilation and potential for aerosol contamination during this procedure would increase danger for patient and staff alike. On the other hand, ventilating COVID-19 patients in an Operating Room (OR) with AGS is undesirable as it reduces hospital operating capacity, makes nursing more difficult, and the OR is a positive pressure environment. Any aerosol viral particles spilled in an unmodified OR is spread throughout the Theatre complex by positive pressure in the room.

A solution that allows use of a humidified circuit, but without AGS was thus sought to allow the use of ICU bed spaces. Such a solution would need to be a reversible change to the machine, pass the normal System Check, and allow ventilation on a humidified circuit for an extended period without AGS, but with no condensation within the anaesthetic machine. 48 hours was chosen as a reasonable test period for any ‘proof of concept’ prototype.

#### A kludge to the rescue?

It was considered that bridging ports on the Patient Cassette with a section of breathing system tubing might provide a solution by replacing the Volume reflector, shortening the gas pathway, and reducing the opportunity for condensation to form. However, although the tubing selected appeared to fit the cassette ports, and tubing could easily be hidden in the gap where the un-needed Volume Reflector normally sits, the machine failed to pass self-test. The failure was due to one of the connectors abruptly disconnecting during the pressure test. It was concluded that even if we could induce the machine to pass test, the risk of disconnection during ventilation would be too high.

#### A manufactured solution?

Given the failure of the previous attempts to provide a workable solution, local manufacturing companies Kilwell Fibretube and subsequently RAM3D were approached and asked to reverse engineer the Socket. The intention being to make a U-tube which connects the Patient Cassette internal input and output tubing together, in order to bypass the Volume Reflector entirely. This device will be referred to as the *Bridge*.

The key design aspects of the Bridge were that it must be simple to install; limit risk of failure during extended periods of operation; seal tightly on the Cassette silicone seals intended for the Socket; not interfere with normal functioning of the Cassette; and be of a material which is suitable for autoclave or other sterilisation methods.

In this instance 3D printing provides a simple, quick to innovate, and cost-effective method of manufacture. It is important to note that FDM style printers (the most common around the world) are not well suited for this device. The nature of the FDM system means that they will be prone to leakage either during pressure testing (which uses approximately 117cm H_2_O (1.7psi; 11.5kPa) pulses of pressure to check the internal gas pathways); or as a result of fatigue during the extended cyclic loading during operation. There are a number of aspects of the Cassette installation which creates risk of damage to an FDM manufactured part during installation. Furthermore, the poor surface finish of FDM will create issues with the seals and cleaning.

The successful prototype was designed and printed by RAM3D (2020). It was printed on a laser powder bed fusion machine from grade 23 Ti64 (medical grade titanium alloy). The key attributes of using a system such as this are the good surface finish, high relative strength, and a body safe material that is already approved for medical use. Using this system also enabled a much simpler single piece design requiring no further assembly and very minor post printing processing to smooth the area of the seal.

A critical factor of the design are the fine tolerances. Alternative manufacturing methods must also reflect this, as even very small variations cause the seals to leak due to poor fit, or due to misalignment of the Cassette with other gas seals.

#### Prototype Test Results

##### Leak tests

The successful prototype passed System Check when installed on six different Flow-i machines in Lakes DHB’s Theatre complex. The average leak was 42ml/min, well within the <150ml/min allowed. Each machine was then restored to stock and immediately passed System Check normally before being returned to use.

##### Extended Use

A test lung was ventilated for 48 hours under the conditions the machine would experience in ICU. Pooled water appeared in the inspiratory limb of the humidified breathing system after 24 hours, but this was easily decanted back into the water bath. That is an intended behaviour of the Fisher & Paykel humidification system. At 48 hours the test was stopped, the anaesthetic machine stripped down and the anaesthetic machine gas pathways examined. There was no water in the patient Cassette. The Bridge and its seals were dry and there was no water in the Scavenging Exhaust system.

#### Outstanding Issues

There are problems which remain to be addressed in order to optimise patient use. The authors hope that by releasing this document and the device with a CC-BY 4.0 license at an early stage, we can access the ‘wisdom of the crowd’ and collaboratively solve these issues.

1. Scale manufacture is available in New Zealand and near markets from RAM3D. However freight logistics have become a problem due to pandemic shutdowns, so design and production may need to be adapted to suit local manufacturing capabilities. Production devices may be printed from a sterilizable patient safe material and reused, as we have done; or printed from a medical grade plastic for single patient use. Other manufacturers will need to confer with hospital Central Surgical Sterilizing Departments and test materials for suitability.
2. The end-tidal CO2 line water trap is likely to fill relatively quickly and will need decanting. This water may be virally contaminated. Internal contamination is not an issue as the sampled air is HEPA filtered and dried prior to entering the gas bench. Work will need to be done to mitigate the risk this poses to ICU nurses, or the Flow-i will need to be used without its gas bench.
3. The Flow-i internal software is designed around the presence of the Socket and Flow Reflector. This may have unforeseen undesired effects during use.
4. Choice of patient circuit will need to be tested by end users depending on local availability.

## Conclusions

The presented ‘proof of concept’ Bridge device holds the potential to allow conversion of many Maquet Flow-i machines currently under-utilised for their primary purpose into Intensive Care ventilators. It may thus help bridge that period of time while more purpose-built Intensive Care ventilators are manufactured. Local manufacture is available to New Zealand and near markets from RAM3D, but the CAD file is made available and allows the reader to replicate the Bridge locally and investigate manufacture by local engineering firms. Choice of material will depend on local manufacturing capability and available materials. This innovation is CC-BY 4.0 licensed and the authors welcome collaboration and clinical exploration of the modification by the wider Anaesthetics, Intensive Care, and Engineering communities.

## Data Availability

Test information is all in the paper. CAD file is available on GrabCAD.

https://grabcad.com/library/flow-i-bridge-1

